# Topical application of Urolithin A slows intrinsic skin aging and protects from UVB-mediated photodamage: Findings from Randomized Clinical Trials

**DOI:** 10.1101/2023.06.16.23291378

**Authors:** D D’Amico, AM Fouassier, J Faitg, N Hennighausen, M Brandt, D Konstantopoulos, C Rinsch, A Singh

## Abstract

Urolithin A is a gut microbiome derived postbiotic that has been shown to stimulate mitophagy, and improve muscle and mitochondrial health when administered orally to humans. In three separate randomized trials, we have now investigated the effect of topical administration of Urolithin A on skin aging features and on UVB-mediated photodamaged skin. Post-menopausal women with evidence of skin aging such as > Grade 3 wrinkle formation were included in a split-face/arm study design in the first trial (*aging study 1; n=48*), followed by a second larger trial (*aging study 2*; n=108) in middle-aged men and women focusing on wrinkle reduction. Healthy participants were included in the placebo-controlled, randomized UVB-induced trial (*photo-damage trial; n=22*). Participants were randomized to receive topical supplementation with either 0.5% Urolithin A cream or placebo for 8-weeks in a low-dose arm or 1% Urolithin A cream or placebo in the high-dose arm in the *aging study 1*. In the *aging study 2* participants were randomized to receive 1% UA in a day-cream, a night cream and a serum, that were compared to the untreated site. For the *photo-damage trial,* topical patches containing either 0.5% or 1% UA or placebo cream were applied for 24-hours following UVB irradiation. The primary outcome in the *aging study 1* was an impact on biological pathways linked to skin aging in skin biopsies, and an impact on skin barrier function after 8-weeks. Key secondary endpoints were a change in facial wrinkle appearance (crow’s feet area). The *aging study 2* focused on wrinkle reduction as a primary outcome. In the UVB-mediated *photo-damage study*, the primary read-out was the change in erythema after application. Molecular analyses were conducted on skin biopsies and using *ex-vivo* systems to investigate the mechanism of action mediating skin protective effects of Urolithin A. In the *aging study 1*, Urolithin A at 1% significantly up-regulated collagen synthesis pathways in human skin biopsies and led to a decrease in wrinkle depth on facial wrinkles. The lower dose had no significant impact. There was no change on skin barrier function with both doses suggesting maintenance of a healthy skin barrier function. In *aging study 2*, topical application of Urolithin A at the 1% dose in different formulations (day-cream, night cream and serum) led to significant wrinkle reduction compared to the untreated side, confirming the previous findings. Skin hydration was improved significantly as well. In the third trial, investigating impact on photodamaged skin, Urolithin A application led to a significant decrease in UV-induced erythema (∼14%) compared to the untreated area, while placebo and lower dose UA cream’s showed no benefits. Urolithin A topical administration was safe and well-tolerated in all studies. UA also inhibited collagen degrading and pro-inflammatory pathways and up-regulated gene expression of biomarkers linked to induction of mitophagy and autophagy in human skin cells. Taken together, these clinical studies support the topical use of Urolithin A to manage and prolong skin health longevity by acting at the cellular level, supporting collagen structure, reducing wrinkle appearance and protecting against photoaging.

The studies are registered in clinicaltrials.gov as: NCT05300984; NCT05473832; NCT05300542

## Introduction

Skin is the largest organ in the body and serves key functions as a barrier against injuries, UV light and pathogens. Chronological aging brings changes in skin appearance and a decline in its protective functions. This is further accentuated following exposure to UV radiation, by a process called photoaging^1^. The most common signs of aging skin are the appearance of wrinkles, skin thinning, hyperpigmentation, sagginess, and dryness. This derives from a combination of biological changes, including a decline in quantity and quality of structural components of the skin, such as collagen and elastin, as well as dysfunctions in cellular bioenergetics. Indeed, most of the energy to support skin cell heath is derived from mitochondria, making mitochondrial dysfunction a key hallmark of skin aging ^2, 3^. Finally, immune cells located in the skin are impacted by the aging process. This reduces the ability of the skin to cope with excessive inflammation by external agents and significantly contributes to the emergence of photoaging ^4^.

Current approaches to counteract skin aging are limited and primarily include retinol or retinoid-based solutions. They are effective, for example, at wrinkle reduction, but do not have the best safety and tolerability profile and are controversial in the management of photoaging^5, 6^. There is a need for new innovative natural bioactives for improving skin health and longevity, that have a dual benefit profile, impacting both intrinsic and extrinsic aging, without irritations observed with retinol and retinoids. Urolithin A (UA) is a food metabolite, and a postbiotic of the gut microbiome that has been shown to both improve mitochondrial health and reduce age-associated inflammation, i.e. inflamm-aging. UA increases mitophagy, the process of recycling faulty mitochondria, and enhances biomarkers associated with better mitochondrial function in several models of aging and age-associated conditions ^7^. In humans, oral supplementation of UA was shown to be safe, improve mitochondrial function, and improve muscle strength and endurance in clinical studies in middle-aged and elderly subjects ^8, 9^.

Topical application of UA represents a promising strategy to support skin health and extend skin longevity through its combined effect on multiple hallmarks of skin aging. We performed a systematic clinical investigation of the benefits of topical application of UA to investigate its safety and impact on clinical readouts of both intrinsic skin aging and photoaging. Finally, we analysed skin biopsies and human cell models to identify molecular mechanisms associated with UA’s protective effect on the skin tissue.

## Methods

### Subject recruitment

All the studies were approved by an independent institutional review board (Institutional Review Board, proderm GmbH, Kiebitzweg 2, D-22869 Schenefeld, Germany). The subjects were recruited at the study site for each of the studies (SGS Proderm, Hamburg, Germany). The suitability of each subject was evaluated according to the various inclusion and exclusion criteria. Subjects gave written informed consent to participate in the study and to come to the scheduled visits. In the first aging study (*aging study* 1) only female participants from 50 to 75 years of age with visible wrinkle in the face (grade 3 to 6 according to wrinkle severity scale) were included. In the second aging study (*aging study* 2), both males and females from 40 to 65 years of age were recruited with similar criteria as the first study. Subjects had healthy skin in the test areas. Prior/concomitant diagnoses and prior/concomitant therapies relevant for the study were recorded. Main exclusion criteria were any documented allergies to cosmetic products and/or ingredients, skin care and/or skin cleansing products and use of any topical medication at the test area within the last 3 days prior to the start of the study. For the *photodamage study*, both females and male participants from 18 to 65 years of age were included. Subjects had uniform skin color and no erythema or dark pigmentation in the test area prior to the start of the study. The skin type of the enrolled subjects was classified by use of colorimetric skin type classification.

#### Randomization and instructions of use

The skin aging studies both followed the split-face study design, i.e., each participant was randomly assigned to the test products to apply on either the left or the right side of the face and arm in case of the first study where skin biopsies were collected. All studies were controlled, randomized studies with the first aging and UVB erythema trial also being double-blind, placebo-controlled studies. The second aging trial utilized a split-face design with a treated and an untreated side for comparison. The tested products were the following for the both the *aging study 1* and the photodamage study: a moisturizing skin base cream containing standardized excipients (vehicle) and for the (active) arms containing either 0.5 or 1% UA (Mitopure^TM^) added in the base cream. The products tested in the *aging study 2* contained 1% UA (Mitopure^TM^) in a day-cream formulation, a night-cream formulation and a serum product. A study site representative assigned test products so that both the subjects and the investigators were blinded to the test article identification. The information containing the assignment of products to labels was kept at the study site by a person not involved in the investigations and analysis of data during the study. Statistical analyses plan was finalized before unblinding. Afterwards the envelope with the assignment of codes to treatments was opened to unblind the study for analysis. A study technician demonstrated to study participants the correct amount of the test product to be applied during the first application. The test materials were applied twice daily in the morning and evening for 8-weeks by the subjects at home according to the application training in the first aging study, whereas in the second trial subjects applied it either in the day (for day-cream) or the evening (for night-cream), or twice daily in the morning and evening for serum. The subjects were instructed to use the test product every day.

**Skin barrier function** was quantified as transepidermal water loss (TEWL) and was measured via a TEWAMETER® TM 300 (Courage & Khazaka, Cologne, Germany). TEWL is a non-invasive method to measure the barrier function of the skin and is regarded as a sensitive parameter to quantify skin barrier damage. One measurement per test area and assessment time was recorded. Briefly, water evaporation from the skin was measured by placing cylindric open chamber with two hygrosensors at a defined distance from the skin. The probe was held in place for each measurement for 30 seconds. The values of the last 10 seconds (= 10 values) were averaged as the actual measurement value.

**Skin roughness and wrinkle assessment** was performed either via a DERMATOP blue (Eo Tech SA, Marcoussis, France) *in aging study 2* or via standardized, computer-controlled facial photography with a 24 Megapixel Nikon camera (Colorface®, Newtone Technologies, Lyon, France) *in aging study 1*. The anti-wrinkle efficacy of each treatment code was assessed in the periorbital regions by investigating the three-dimensional structure of the wrinkles. These assessments were conducted at 2 and 8 weeks in the first aging study and at 2, 4 and 8 weeks in the second study. Using phase-shifted and gray-coded measurements of human skin the three-dimensional surface structure of the investigated skin site was captured. The measuring principle is based on digital fringe projection. The fringes that are projected under a defined triangulation angle onto the surface of the measured target with a sinus-like intensity of brightness are detected with a CCD camera. The three-dimensional skin surface profile is calculated from the position of the fringes in combination with the gray values of each pixel. From the captured three-dimensional structure, roughness parameters are calculated. Parameters: Rz and Ra are chosen, representing mainly the rough structure (Rz) or the finer skin structure (Ra). A decrease in the roughness parameters Rz and Ra corresponds to a decrease in the degree of skin roughness. 1 measurement per test area and assessment time was performed.

**Skin hydration assessment** was measured via the CORNEOMETER CM 825 (Courage & Khazaka, Cologne, Germany). The measurement of stratum corneum hydration was performed by the electrical capacitance method with the corneometer. The measuring principle is based on changes in the capacitance of the measuring head, functioning as a capacitor. Between the conductors consisting of gold, an electrical field is built. By these means, the dielectricity of the upper skin layer is measured. Because the dielectricity varies as a function of the skin’s water content, the stratum corneum hydration can be measured. Five measurements per test area and assessment time were performed and the average recording used for statistical analysis.

### Skin biopsy excision (from *aging study 1*)

The room and biopsy area (dorsal forearms) were disinfected prior to the procedure with the disinfectant spray (e.g. Octenisept®). A local anesthetic (e.g. Skandicain® 1%) was administered to the biopsy site subcutaneously using a sterile syringe, fitted with a suitable sterile needle, to the biopsy area prior to excision of the biopsy. Punch biopsies (3-mm) were be sampled by a physician using a punch biopsy instrument. The biopsy specimen contained the epidermis and the dermis. Each biopsy sample was then appropriately labelled (with the study and subject number). Following the biopsy excision, the wound was closed with SteriStrips® and covered with a protective dressing. The subjects were supplied with waterproof dressings for covering the wounds after skin biopsies were taken. The wounds had to be kept dry until the wounds healed. Instructions on changing the dressing and spare dressings were provided to the subjects. In order to reduce the risk of hyperpigmentation the participant’s undergoing skin biopsy procedure were instructed to avoid sunlight at the test areas after conduct of the study and/or to use a sun protection cream. Biopsies were collected at baseline and end of the study.

### MED dose calculation, UVB erythema induction and assessments

A solar simulator multiport 300 W (SOLAR Light Co, Philadelphia, USA) lamp equipped with appropriate filtration was used to irradiate the skin test areas with the requested irradiation intensity on spots with a diameter of 1 cm. A single irradiation per test area was performed. Test areas were outlined with a skin marker on the backs of the subjects. Thereafter, baseline measurements were performed on all test areas. After that six spots for minimal erythema dose (MED) determination were irradiated on the back with a diameter of 0.9 cm each. The UVB dose was increased from spot to spot by an increment of 25 %. An untreated irradiated control area served as a negative control. After irradiation, the test products were applied using an occlusive patch to the irradiated spots post irradiation. 23 ± 2 hours after irradiation, subjects returned to the study site and product topical patches were removed. Residues of the test products were carefully removed with a dry soft paper tissue. The MED dose and a change in skin redness (erythema score) from baseline was recorded via a CHROMAMETER CR 400 (Minolta, Device D-Langenhagen, Germany). The Chromameter defines the measured color in the L*a*b* color coordinate system. The L*-value defines the brightness. The color is defined by the parameters a* (red-green axis; negative a*-value green, positive a*-value red) and b* (blue-yellow axis, negative b*-value blue, positive b*-value yellow). The a* value correlates well with visual assessments of skin redness (erythema). a* is a measure for erythema. An increase in the a*-value corresponds to an increase in the degree of skin redness. A trained grader also assessed the skin erythema for each group of irradiation dose of the different test areas, according to the following scale: -2=marked increase in redness compared to negative control (untreated); -1 =slight increase in redness compared to negative control (untreated); 0=no difference in redness compared to negative control (untreated) (no effect); 1=slight decrease in redness compared to negative control (untreated); 2=marked decrease in redness compared to negative control (untreated); 3=complete suppression of redness.

### Adverse reactions recording

All ICH GCP guidelines of recording of adverse reactions (ARs) were followed. For all the clinical studies all ARs were assessed by the study physician for a causal relationship between the test material and the adverse event (AE) was established. All adverse reactions (excluding those parameters being scored as part of the protocol) were documented in the study records. ARs were recorded from study enrolment of the subject in a study to until 5 days following last administration of test product.

#### RNA-seq

Skin tissue samples were disrupted in a TissueLyser and RNA extraction performed using an automated QiaSymphony extraction robot and the QIAsymphony RNA Kit. Strand-specific cDNA library preparation was performed by purification of poly-A containing mRNA molecules, mRNA fragmentation, random primed cDNA synthesis (strand-specific), adapter ligation and adapter specific PCR amplification. RNA-seq run was performed using the NovaSeq6000 using S4 flowcells with 2×150bp. Quality Control (QC) and preprocessing of raw reads was performed by an in-house QC pipeline. The pipeline utilizes fastP version 0.23.2 [Chen, 2018], for quality control, quality trimming, and adapter clipping procedures. Reads with a minimum PHRED score of 15, a minimum length of 15 bp, and a minimum unqualified percent limit of 40 (40%) were retained for downstream analysis. QC reports were summarized by MultiQC version 1.12 [Ewels et al., 2016]. Quality-processed read pairs were mapped against the Ensembl human reference genome GRCh38.99, using a transcriptome annotation GTF file (version GRCh38.105). In particular, an in-house RNA-sequencing alignment pipeline that utilizes the STAR algorithm version 2.6.1c [Dobin et al., 2013] was applied. Read counting was performed on the gene-level, using htseq-count [Putri, 2022]. To inspect the alignment results, MultiQC was used accordingly. Read counts were normalized to account for inherent compositional and technical biases using “median of ratios” [Anders and Huber, 2010] method from DESeq2 R package [Love, 2014]. Differential expression analysis was applied for Mitopure condition, controlling by vehicle condition, and by comparing samples between two visits (T2 vs T1), adjusting the DESeq2 design formula accordingly (design = ∼ Treatment + visit + Treatment: visit). Before DEA, a pre-filtering step was applied to remove low abundance mRNA measurements, by excluding genes with less than 10 total counts across samples. Additionally, an independent filtering procedure of DESeq2 was enabled, to filter out genes with very low counts that are unlikely to show significant alterations in gene expression. Gene symbols (HGNC nomenclature), descriptions, and biotypes were matched to Ensembl GTF ids by using the R package biomaRt. v. 2.46.3 [Durinck et al., 2009]. P-values were corrected for multiple-testing using the Benjamini and Hochberg (BH) method [Benjamini and Hochberg, 1995].

#### Gene Set Enrichment Analysis

Gene Set Enrichment Analysis (GSEA) ^10^ was performed by using the R package ClusterProfiler^11^. All processed and filtered genes (genes with at least 10 total counts across samples having an applicable p-adjusted value) were ranked by the comparison’s log2 fold change, and the enrichment of a given gene set among “activated”- or-“suppressed” genes was statistically tested. Gene sets from ontologies within Gene Ontology (GO) ^12^ Biological Processes (BPs) were tested. The minimum and maximum gene set sizes were set to 10 and 500, respectively. The resulting p-values were adjusted using the BH method. Enriched terms with a maximum adjusted p-value of 0.05 were considered statistically significant. GSEA significant results were visualized as dot plots using the enrichplot R package ^11^. Expression changes of core enrichment genes of significant GO BP terms for the “Mitopure normalized over vehicle” comparison was further visualized. For the particular analysis, only subjects present in both visits (T1 and T2) and both treatments (Mitopure and vehicle) were considered. Initially, for each treatment, each subject’s T2 expression values were corrected by their corresponding T1 expression. Then, Mitopure expression changes were further corrected by the vehicle expression changes, between the respective subjects. Ratios are referred as “Fold changes over vehicle”. Finally, log2 Fold changes over vehicle were visualized as heatmaps, by utilizing pheatmap R package [Kolde, 2018]. Genes were hierarchically clustered with a euclidean distance metric using complete linkage.

#### Skin aging dataset analysis

A text file depicting gene expression changes with age in skin was retrieved by the supplemental material of a previously published study ^13^. The particular dataset was reanalyzed by GSEA as described above, adjusting “Beta” value which was considered a ranking factor instead of log2 Fold Change. Significant suppressed GO BP terms were intersected with the corresponding significant activated GSEA results of “Mitopure normalized over vehicle” comparison, resulting to one common BP term, “Collagen Fibril Organization”. Common core enrichment genes between the two instances of the particular term were further processed, and visualized as follows: (a) For the RNA-seq dataset, bar plots of “Fold changes over vehicle” (see above for definition) were plotted, while (b) the aging dataset was visualized as bar plots of Beta values.

#### Quantitative real-time PCR (q-RT-QPCR)

RNA from cells and reconstituted epidermis was extracted using TRIzol (Thermo Scientific, 15596026) and then transcribed to cDNA by the QuantiTect Reverse Transcription Kit (Qiagen, 205313) following the manufacturer’s instructions. The q-RT-PCR reactions were performed using the TaqPath ProAmp master mix (Applied biosystems, A30866) and the expression of selected genes was analysed using the Quantstudio 6 flex (Life technologies) and following TaqMan probes listed in Supplementary table 3. All quantitative polymerase chain reaction (PCR) results were presented relative to the mean of housekeeping genes (ΔΔCt method). mRNA levels were normalized over RPLP2 for gene expression for cell and tissue samples.

#### Reconstituted human epidermis

Ten-day-old reconstructed human epidermis (RHE; batch 01015-587) were placed in proprietary maintenance medium (Bioalternatives). Topical application with a moisturizing cream with or without Mitopure at 1% was performed for 24 hours. The RHE were then rinsed in PBS and irradiated with UVB (+UVA) - 850 mJ/cm² (+ 6 J/cm²) using a SOL500 Sun Simulator equipped with an H2 filter (Dr. Hönle, AG). After irradiation, the treatments were renewed and the RHE incubated for 24 hours. In parallel, non-irradiated conditions were performed and kept in the dark during the irradiation time. PGE2 released in the culture supernatants was measured using a specific ELISA kit according to the supplier’s instructions (Enzo Life Sciences, ADI-900-001). All experimental conditions were performed in n=3.

#### Statistical analysis

The mean values of raw data and calculated values were presented in bar charts with 95 % confidence limits for each treatment and assessment time. A significance level of 0.05 (alpha) was chosen for statistical analysis. Due to the proof-of-concept character of the study, no adjustment for multiplicity was done. Pairwise comparison of treatments was done by paired t-test on differences to baseline for each assessment time and treatment group. For visual and imaging data pairwise comparison of treatments was done by Wilcoxon signed rank test on raw data. The computation of the statistical data was carried out with commercially available statistics software (SAS for Windows).

## Results

### Demographics

For the *aging study 1*, a total of 55 participants were screened in the placebo-controlled, first aging study. Of them, n=48 middle aged women subject with mean age of 58.6 ± 6.6 years were randomized (Figure 1) in to two study arms with a split-face/arm study design with a lower dose active group compared to placebo and a second higher dose active group compared to placebo. All participants completed the trial and there were no dropouts. Ethnicity of all participants was Caucasian. In the *aging study 2*, n=108 middle-aged subjects with a mean age of 55.4 ± 6.6 years were randomized using a split face study design with a treatment side compared to an untreated side. All subjects completed the study protocol (Supplementary Figure 1A). There were 29 males (27 %) and 79 females (73 %) in the final analysis of which n=2 (2 %) were Black, 93 (86 %) were Caucasian, n=3 (3 %) East Asian, n=3 (3 %) Hispanic and n=7 (6 %) West Asian ethnicity. In the UVB-mediated erythema trial (placebo-controlled, Photo-damage study), n=22 subjects (10 male (48 %), 11 female (52 %) with mean age 44.0 ± 14.1 years and Caucasian ethnicity) were enrolled. There was one drop-out subject and n=21 subjects completed the study protocol (Supplementary Figure 1B).

**Figure 1.**
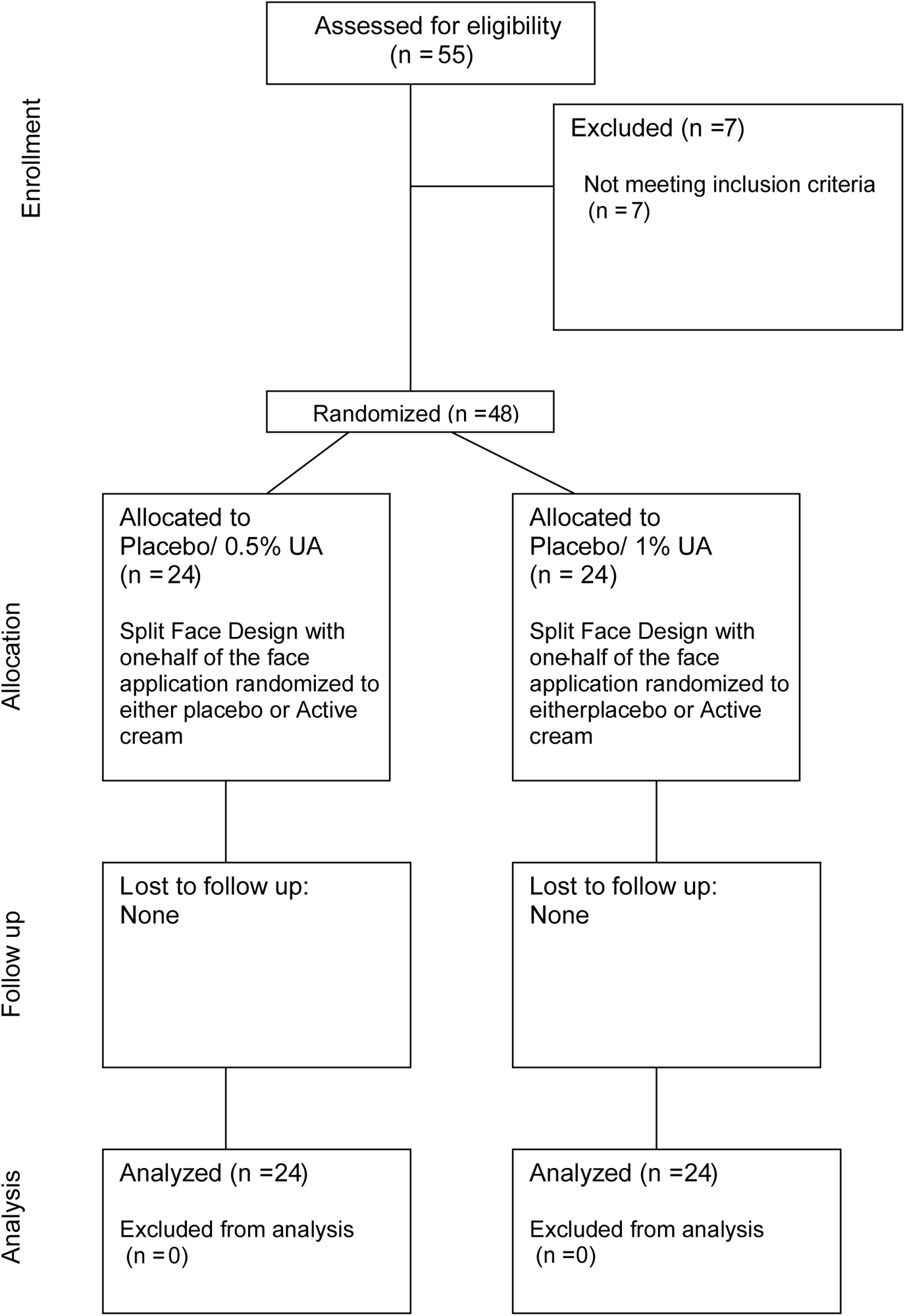
CONSORT Diagram of *Aging Study 1*. 55 Participants were screened in the placebo-controlled, first aging study of which n=48 middle aged women subject with mean age of 58.6 ± 6.6 years were randomized to two study arms with a split-face/arm study design with a lower dose active group compared to placebo and a second higher dose active group compared to placebo. Skin biopsies were conducted on the volar forearm in adjacent areas at the start and end of the 8-week intervention. All participants completed the trial and were included in the final analysis.

### Safety

In the *aging study 1*, no adverse reactions with any relationship to study products were reported by any subject. In the *aging study 2*, four adverse reactions occurred across three subjects as varying forms of skin irritation, including erythema, papules, itching and burning. Of the adverse reactions documented during the conduct of this study, all were classified as being of mild severity (Supplementary Table 1). In all cases, subjects remained in the study and recovered without sequelae. In the *photodamage trial*, two adverse reactions occurred in the form of skin irritation with erythema in one subject. The reactions were of mild severity and resolved without sequelae.

### Wrinkle reduction with 1% topical UA application

In the *aging study 1*, wrinkle reduction was measured after 2, and 8-weeks of test treatment application (Fig. 2A). The UA 1% treatment group showed a significant reduction in wrinkle depth at 8-weeks (p=0.04; Figure 2B) compared to the placebo treated side. UA at 0.5% did not showed any significant effect compared to the Placebo (Supplementary Table 2). In the *aging study 2*, UA at 1% was found to have a significant impact on reducing wrinkles (finer and rougher skin structures computed as Ra and Rz, respectively) with the effect already manifesting after 2 weeks of application and maintained with 4 and 8-weeks of application, compared to the untreated side (Supplementary Table 2). The effect was independent of timing of application and matrix of the product applied, as the three products (day cream, night cream and serum) containing 1% UA showed similar wrinkle reducing benefits (Figure 2C, day-cream; Figure 2D, night-cream; data for serum are shown in Supplementary Table 3).

**Figure 2.**
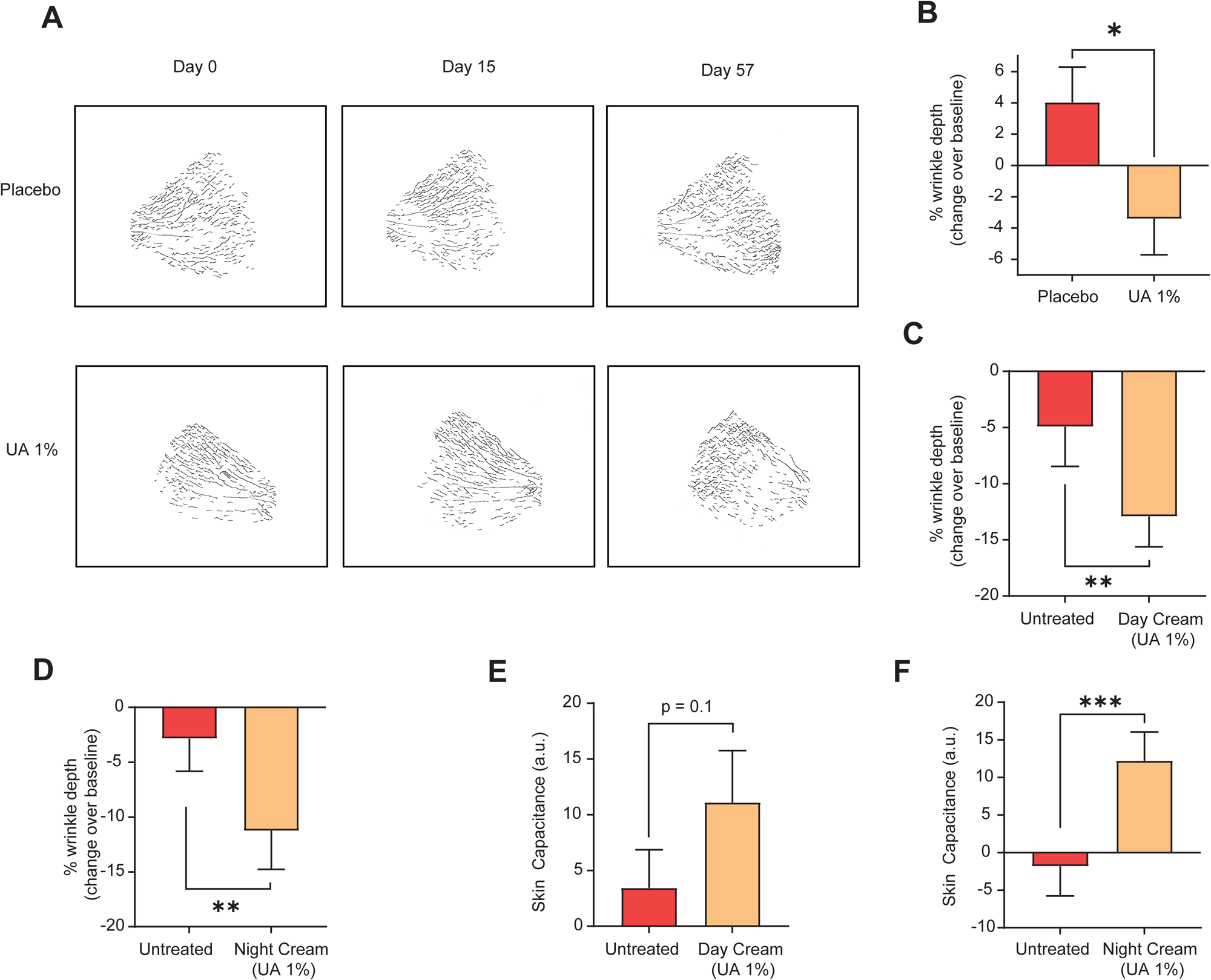
Topical application of UA leads to wrinkle reduction effect. UA 1% topically applied in a basic skin cream (**A**) in the *aging study 1* or in a more defined matrix of day (**B**) and night (**C**) skin cream formulations from the *aging study 2*, exhibited significant anti-wrinkle effects in middle-aged subjects with moderate facial wrinkles after 8-weeks of topical application (**p<0.05**). Skin barrier function remained healthy and unchanged over the course of the *aging study 1* (**D**). Skin hydration was improved upon topical application of different UA containing creams in *aging study 2* (**E, F**).

### Impact on Skin barrier and skin hydration

All subjects entering the skin aging studies had a healthy skin barrier and topical UA application showed no negative impact on skin barrier, with function remaining unchanged(Figure 2D). Daily application of the creams (day, night creams and serum) containing 1% UA significantly improved skin hydration after 2-weeks of use in comparison to pre-test treatment level. The observed improved skin hydration levels, as measured by skin capacitance, were maintained also after 8 weeks of application in comparison to the untreated side (Figure 2E, F, Supplementary Table 4).

### Supportive biomarkers

We investigated biological pathways impacted by topical UA application for a period of 8 weeks by performing RNA-seq transcriptomics of forearm skin biopsies from the *aging study I*. Gene set enrichment analysis (GSEA) was applied to identify significant pathways changed by UA comparing samples at the study end to baseline and normalizing results over vehicle. “Collagen fibril organization” was the molecular pathway most significantly activated by UA (Fig. 3A and Suppl. Fig. 3A), with UA application also reducing gene sets related to chemokine response (Fig. 3A). We further studied which UA regulated pathways are involved in the natural skin aging process. Analysing a publicly available skin aging clinical study ^13^, the “Collagen fibril organization” was a common signature, both induced by UA and downregulated with skin aging (Suppl. Fig. 3B). This gene set includes collagen genes encoding for fibrillar type I and III collagen (Fig. 3B), known to decline in skin with chronological aging ^14^, photo-aging and smoking ^15^. UA also upregulated age-associated decline of genes involved in skin homeostasis. Among them, *EMILIN-1*, regulating 3D structure and elasticity ^16^ and *AEBP*, encoding for a collagen-interacting protein required for optimal tissue repair ^17^ (Fig. 3B). These data provide mechanistic insight into the positive impact of UA on wrinkle reduction, a process that is driven by impaired collagen production and organization in the skin. The GSEA results reveals insights on the impact of UA on the dermis, the skin layer producing and secreting collagen.

**Figure 3.**
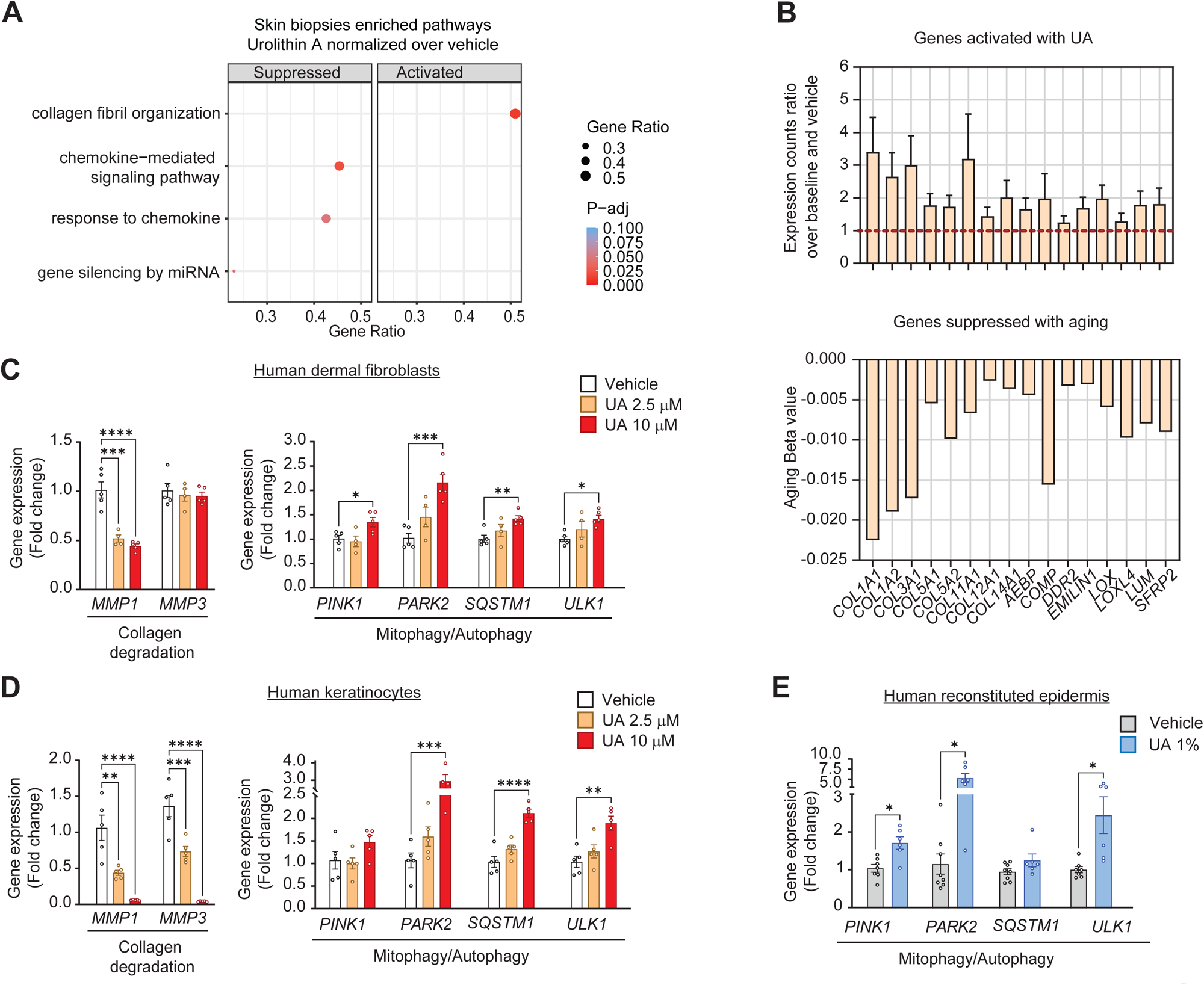
UA positively impacts biomarkers associated with improved skin health. (**A**) GSEA results from RNA-seq in skin biopsies from *aging study 1* showing the most significant Gene Ontology Biological Processes (GO-BP) pathways that are significantly enriched in UA compared to vehicle. Terms with positive and negative normalized enrichment scores (NES), are categorized as “activated” and “suppressed”, respectively. Pathways are sorted by gene ratio, dot sizes scaled by the number of core enriched genes, and dot color reflects the statistical significance of the enrichment (Benjamini-Hochberg-adjusted p-value, P-adj). (**B**) Bar plot showing increased expression in core genes of the “Collagen fibril organization” pathway in skin biopsies exposed to UA compared to the vehicle group (top). Bar plot of Aging Beta value for same genes derived from the skin aging trial (bottom). Only common core enriched genes between UA and skin aging dataset are shown. (**C-D**) Expression of genes encoding for collagen degrading enzymes (*MMP1, MMP3*) and autophagy/mitophagy proteins (*PINK1, PARK2, SQSTM1, ULK1*) in human dermal fibroblasts (C) and human keratinocytes (D) treated for 24 hours with DMSO or the indicated doses of UA. (**E**) Expression of genes encoding for autophagy/mitophagy proteins (*PINK1, PARK2, SQSTM1, ULK1*) in human reconstituted epidermis after daily application for 72 hours of Vehicle or UA 1% containing moisturizers. Data expressed as Fold change over Vehicle +/- sem. *p< 0.05; **P < 0.01; ***P < 0.005; ***P < 0.001 One-Way ANOVA.

To better understand the mechanisms of action of UA in dermal cells, we treated primary human dermal fibroblasts with UA at either 2.5 μM or 10 μM, or with vehicle, for 24 hours. Surprisingly, this short-term exposure to UA did not alter collagen gene expression (Suppl. Fig. 3C). However, it suppressed the expression of the gene involved in collagen degradation, Matrix Metalloproteinases (*MMP1*) (Fig. 3C). Moreover, UA induced a dose-dependent increase in the expression of the mitophagy genes *PARKIN* and *PINK1* and the autophagy genes *MAP1LC3* and *ULK1*, compared to vehicle-treated cells (Fig. 3C).

Human primary keratinocytes treated with UA or vehicle for 24 hours showed a similar gene expression signature as dermal cells. UA led to a significant downregulation of both *MMP1* and the other key collagen degrading enzyme, *MMP3* (Fig. 3D), and it upregulated autophagy and mitophagy gene expression, as in dermal fibroblasts (Fig. 3D). Finally, we employed a model of 3D reconstituted human epidermis (RHE) applied daily with the UA 1% moisturizing cream for 3 days. No change was observed in *MMP1* and *MMP3* gene expression (Suppl. Fig 3D), however, consistent with the data in primary cells, an early activation of mitophagy and autophagy genes in RHE applied with UA compared to Vehicle in the RHE model (Fig. 3E).

## Photodamage study

### Efficacy

In the *photodamage study* all erythema (a*)-values measured 24 hours after irradiation were higher than at baseline for all treatment codes, indicating the development of a sunburn on all the tested areas. The chromameter measurements showed significantly less skin redness (erythema) for 1 % UA treated group after UV exposure at the 1.6MED dose in comparison to the respective untreated area (-13%, p=0.02; Figure 4A), whereas neither the lower concentration with 0.5% (data not shown) nor the placebo showed a significant effect compared to the untreated control.

**Figure 4.**
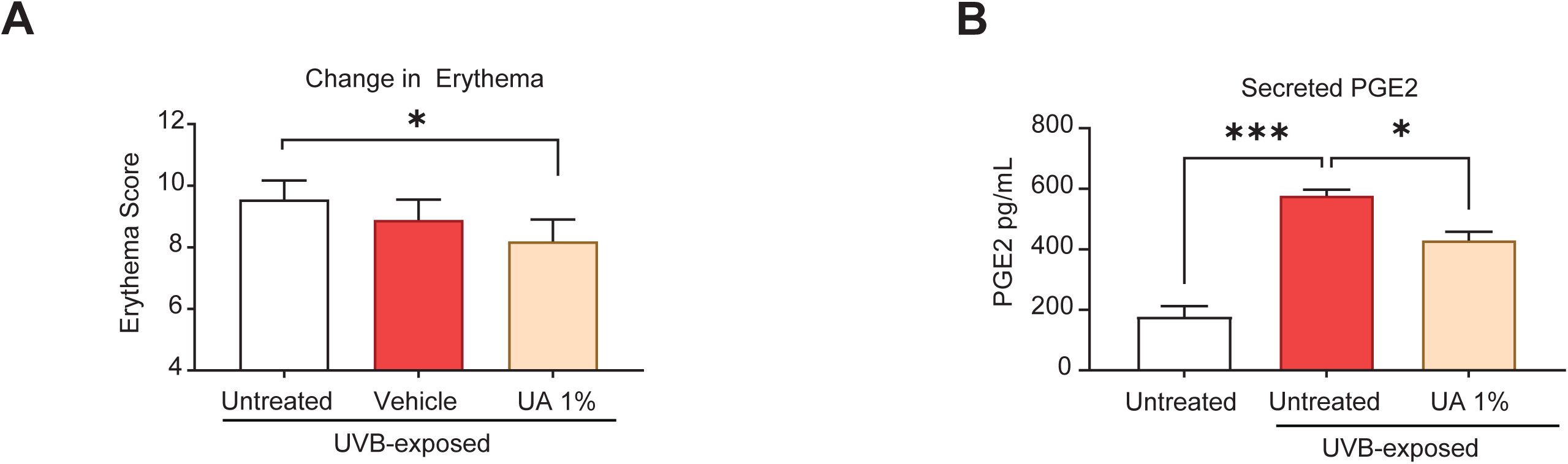
(**A**). (**B**) Levels of prostaglandin E2 (PGE2) secreted in the media of 3D reconstituted human epidermis in the indicated experimental conditions. Data expressed as Fold change over Vehicle +/- sem. *p< 0.05; ***P < 0.005. One-Way ANOVA.

### Supportive Biomarkers

The clinical data from the *photodamage study* supports a protective effect of UA against UV irradiation induced skin damage. We used the RHE to investigate whether this was associated with reduction in the reduction of detrimental inflammation generated by UV irradiation. RHE were applied topically with either the basal topical formulation or a formulation containing UA at 1%. After 24 hours of incubation, RHE tissues were exposed to UVB irradiation and then incubated with the same topical formulations for an additional 24 hours. A group of RHE were left untreated and non-irradiated as negative controls. We measured the concentration of prostaglandin G2 (PGE2) in the culture supernatant, as biomarker of inflammation. Irradiated RHE showed significant increase in PGE2 release compared to non-irradiated tissues (Fig. 4B). UA 1% application of irradiated RHE significantly reduced PGE2 levels, indicating an anti-inflammatory action or the active (Fig. 4B).

## Discussion

Measures to protect against skin aging are essential for maintaining healthy, youthful-looking skin and reducing the risk of skin damage and diseases. In this set of clinical studies, we showed that topical application of skincare products containing UA significantly counteract both natural intrinsic cellular aging and extrinsic photoaging. We performed three complementary randomized clinical studies to document both safety and efficacy of topically applied UA on the skin of healthy adult subjects. Safety was confirmed by a lack of any serious side effects in all participants. In addition, a human-repeat insult patch test conducted (in n=109 subjects, NCT05079607) prior to the start of these randomized trials revealed no irritant or sensitization potential of UA on human skin (data not shown).

Efficacy of the active UA was observed both when applied alone in a basal topical cream formulation (*aging study 1*) and when applied in a day, night cream or serum matrix with a calibrated dose of UA (*aging study 2*). Skin wrinkles are a hallmark of skin aging, especially on facial skin and are known to contribute to the overall rate of aging by inducing oxidative stress and via skin cells that acquire a pro-inflammatory state. Skin wrinkles in the crow’s feet area around the eyes were significantly reduced by an 8-week daily application of UA in multiple skin cream matrices (basal cream formulation, a day-cream, a night cream and serum all containing the same dose of 1% UA). This effect was visible as early as 2-weeks after topical application in all the studies and with the different formulations (Supplementary Table xx). At the molecular level, a hallmark of skin aging is the decline in content and organization of collagen and the extracellular matrix, which provides structural support within the skin tissue. Our data from unbiased skin biopsy RNA-sequencing, showed that topical application of urolithin A increases the expression of the same collagen proteins down-regulated upon natural aging. Further mechanistic analysis in primary human cells and in 3D human skin models revealed that short-term exposure to UA increased mitophagy and autophagy related genes. This is consistent with the molecular signature previously shown in several preclinical models and in human clinical studies in the skeletal muscle. In the same skin models, UA also significantly reduced expression of enzymes causing collagen degradation.

In addition to the protective effects of UA on intrinsic cellular aging, we provide evidence that UA topical application counteracts photoaging. The exposure of excessive UVB radiation from sun is linked to increased skin oxidative stress and an acceleration of skin tissue aging. UA was already shown to protect against UVB-induced photoaging in 2D human dermal fibroblasts, through the activation of mitophagy ^18^. In the current report, we demonstrate for the first time in humans that topically applied UA, in a randomized, double-blind trial, protects against UVB exposed skin, with effects observed following a little as 24-hours after UV exposure that was equivalent to a moderate sunburn. We demonstrated the active UA doses required to significantly reduce UVB irradiation induced erythema formation. Employing a 3D human skin model to investigate mechanistic benefits of UA, we observed a reduced production of prostaglandin E2 (PGE2), a pro-inflammatory molecule induced by UV irradiation.

Compared to UA, other actives employed to counter skin aging have either a less favourable safety profile or more limited range of beneficial effects. Retinol is a commonly used active known to increase collagen production, reduce fine lines, wrinkles and to improve photoaging ^5, 19^. However, retinol increases cell turnover in skin, which results in peeling and dry and flaky effect on skin. It has been shown to cause a dose dependent irritation and erythema with repeated use^20^. In addition, retinol gets degraded on exposure to light and air ^21^.

There are limited interventions to counteract the effects of both skin natural aging that leads to wrinkle reduction and photoaging, without undesirable side effects. In these randomized clinical studies, across ∼180 subjects, topical application of the natural compound UA was shown to be safe and achieved statistically significant improvement in the signs of both intrinsic aging (wrinkle reduction) and extrinsic aging, i.e., UVB-induced photodamage (erythema) in healthy subjects. The benefits were associated with the modulation of collagen and mitochondrial related pathways that contribute to the skin aging processes. Taken together, these results show that UA is a safe and effective natural intervention to counteract skin aging.

## Author Contributions

A.S., C.R. and D.D. contributed to the design of the study. A.S. oversaw the clinical study conduct and operations. N.H. and M.B. coordinated on-site clinical study execution and sample collection as Principal investigators; D.D. oversaw the biomarkers analyses. A.M.F. performed sample processing, and cell-based experiments. A.F., D.D. analyzed the ex-vivo data. D.K., performed all bioinformatic analyses. D.D., J.F., C.R., and A.S. interpreted the data and wrote the manuscript, with the help of the other co-authors. All authors reviewed the manuscript.

## Data Availability

All data produced in the present study are available upon reasonable request to the authors

## Acknowledgement

The authors would like to thank all the study participants for their involvement in the different clinical studies.

## Supplementary Figure Legends

**Supplementary Figure 1.** CONSORT Flow Diagram for (A) *Aging Study 2* and (B) UVB-induced Erythema, *Photodamage Trial*

**Supplementary Figure 2. Impact of Mitopure on biomarkers associated with improved skin health**

**(A)** Heatmap of log2 fold changes from core enriched genes of the “Collagen fibril organization” pathway comparing end of study to baseline, in UA skin biopsies after normalization over vehicle controls. Only subjects present in both visits and both treatments are considered in visualization. Rows (genes) were hierarchically clustered with a Euclidean distance metric using complete linkage. (**B**) Normalized enrichment score (NES) of “Collagen fibril organization” pathway for UA normalized over vehicle comparison and for the “aging” dataset. The significance level of the BP enrichment for each comparison/dataset is denoted accordingly. (**C**) Expression of the gene encoding for Collagen Type I Alpha 1 Chain (*COL1A1*) in human dermal fibroblasts treated for 24 hours with DMSO or the indicated doses of Mitopure. (**D**) Expression of genes encoding for collagen degrading enzymes (*MMP1, MMP3*) in human reconstituted epidermis after daily application for 72 hours of Vehicle or Mitopure 1% containing moisturizer. Data expressed as Fold change over Vehicle +/- sem.

