## Supplementary figures and images for "Topical application of Urolithin A slows intrinsic skin aging and protects from UVB-mediated photodamage: Findings from Randomized Clinical Trials"

### Supplementary Figures

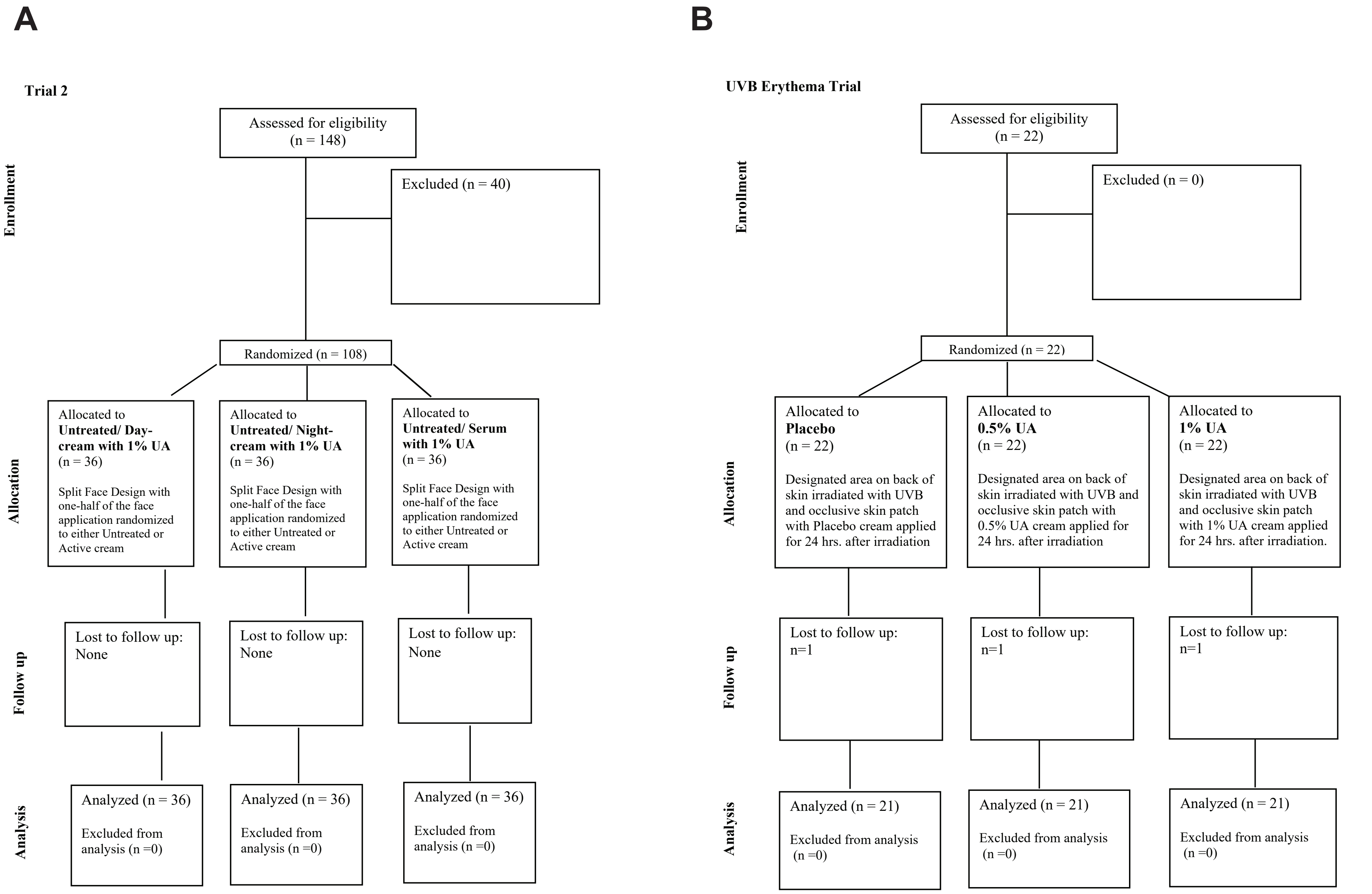


**Supplementary Figure 1**

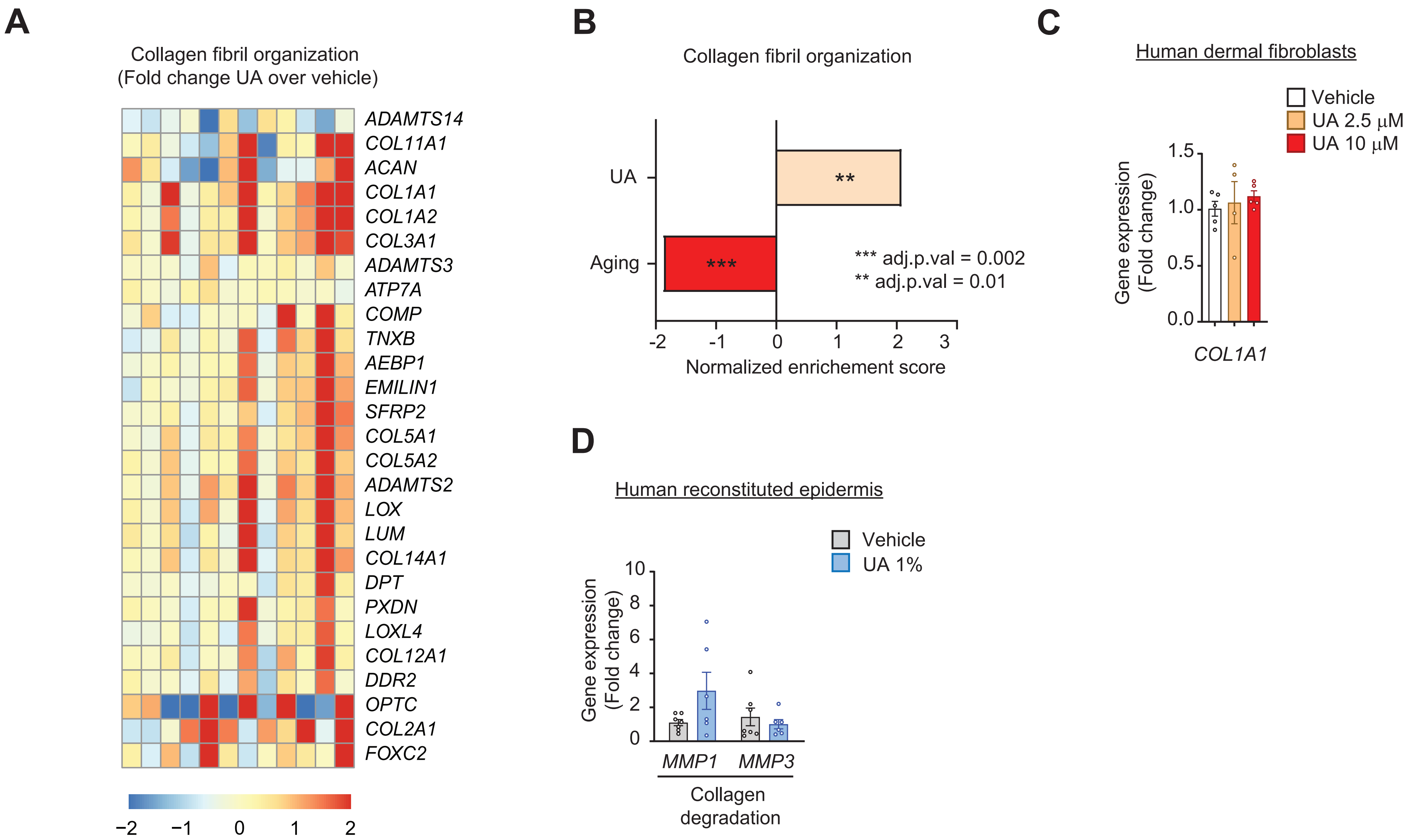


**Supplementary Figure 2**
