## Supplementary Table 4 for "Topical application of Urolithin A slows intrinsic skin aging and protects from UVB-mediated photodamage: Findings from Randomized Clinical Trials"

Skin capacitance as a function of skin hydration was measured on day 1 (baseline, before application of the test product), as well as after 2 weeks, 4 weeks and 8 weeks of product application.

Skin capacitance was measured by Corneometer [a.u.] An increase in Corneometer values correspond to a skin-moisturizing effect.

Mean values, standard deviations, treatment comparison on differences to baseline, and time comparison on raw data for **day-cream, night-cream and serum** containing 1% UA

### A) Day-Cream containing 1% UA

| | | | Mean Values ± SD | | p-Values | | | |
|---|---|---|---|---|---|---|---|---|
| | | | | | Time Comp. | | | Treat. Comp. |
| Time | Code | n | Raw Data | Diff. to BL | vs. BL | vs. D15 | vs. D29 | vs. A |
| BL | A | 36 | 55.50 ± 11.38 | -- | -- | -- | -- | -- |
| | B | 36 | 55.38 ± 11.86 | -- | -- | -- | -- | -- |
| D15 | A | 35 | 55.31 ± 12.45 | -0.31 ± 9.71 | 0.849 | -- | -- | -- |
| | B | 35 | 61.30 ± 10.98 | 5.92 ± 10.34 | **0.002** | -- | -- | **0.004** |
| D29 | A | 35 | 60.37 ± 11.48 | 4.68 ± 9.32 | **0.005** | **<0.001** | -- | -- |
| | B | 35 | 64.20 ± 10.47 | 8.56 ± 10.24 | **<0.001** | **0.038** | -- | **0.028** |
| D57 | A | 36 | 56.33 ± 11.83 | 0.83 ± 10.92 | 0.650 | 0.257 | **0.005** | -- |
| | B | 36 | 59.53 ± 11.13 | 4.15 ± 11.74 | **0.041** | 0.356 | **0.010** | 0.151 |

**bold p-Value: significant (p≤0.05)**

**A:** Untreated side of the face**; B:** 1% UA Day-cream treated side of the face

### B) Night-Cream containing 1% UA

| | | | Mean Values ± SD | | p-Values | | | |
|---|---|---|---|---|---|---|---|---|
| | | | | | Time Comp. | | | Treat. Comp. |
| Time | Code | n | Raw Data | Diff. to BL | vs. BL | vs. D15 | vs. D29 | vs. A |
| BL | A | 36 | 61.09 ± 10.29 | -- | -- | -- | -- | -- |
| | C | 36 | 60.74 ± 10.55 | -- | -- | -- | -- | -- |
| D15 | A | 32 | 61.71 ± 9.57 | 1.02 ± 9.02 | 0.527 | -- | -- | -- |
| | C | 32 | 68.64 ± 7.40 | 8.21 ± 9.94 | **<0.001** | -- | -- | **<0.001** |

| | | | Mean Values ± SD | | p-Values | | | |
|---|---|---|---|---|---|---|---|---|
| | | | | | Time Comp. | | | Treat. Comp. |
| Time | Code | n | Raw Data | Diff. to BL | vs. BL | vs. D15 | vs. D29 | vs. A |
| D29 | A | 36 | 63.92 ± 10.57 | 2.82 ± 10.42 | 0.113 | 0.160 | -- | -- |
| | C | 36 | 67.28 ± 7.88 | 6.54 ± 11.48 | **0.002** | 0.110 | -- | **0.016** |
| D57 | A | 32 | 58.71 ± 13.26 | -1.92 ± 12.86 | 0.406 | 0.164 | **0.005** | -- |
| | C | 32 | 66.39 ± 10.33 | 5.81 ± 12.30 | **0.012** | 0.353 | 0.327 | **<0.001** |
| **bold p-Value: significant (p≤0.05)** | | | | | | | | |

**A:** Untreated side of the face; **C:** 1% UA Night-cream treated side of the face

**C) Serum containing 1% UA**

| | | | Mean Values ± SD | | p-Values | | | |
|---|---|---|---|---|---|---|---|---|
| | | | | | Time Comp. | | | Treat. Comp. |
| Time | Code | n | Raw Data | Diff. to BL | vs. BL | vs. D15 | vs. D29 | vs. A |
| BL | A | 34 | 55.76 ± 10.04 | -- | -- | -- | -- | -- |
| | E | 34 | 54.49 ± 9.62 | -- | -- | -- | -- | -- |
| D15 | A | 32 | 53.28 ± 11.45 | -2.20 ± 9.76 | 0.212 | -- | -- | -- |
| | E | 32 | 65.20 ± 12.03 | 11.14 ± 14.43 | **<0.001** | -- | -- | **<0.001** |
| D29 | A | 33 | 55.68 ± 9.76 | 0.02 ± 9.92 | 0.991 | 0.077 | -- | -- |
| | E | 33 | 62.98 ± 8.70 | 8.70 ± 11.32 | **<0.001** | 0.236 | -- | **<0.001** |
| D57 | A | 34 | 51.82 ± 9.09 | -3.94 ± 10.56 | **0.037** | 0.486 | **0.020** | -- |
| | E | 34 | 63.95 ± 7.99 | 9.46 ± 11.77 | **<0.001** | 0.573 | 0.330 | **<0.001** |
| **bold p-Value: significant (p≤0.05)** | | | | | | | | |

**A:** Untreated side of the face; **E:** 1% UA Serum treated side of the face

BL: Baseline; D15: Day 2-weeks of application; D29: 4 weeks of application; D57: 8 weeks of application
