## Supplementary Table 3 for "Topical application of Urolithin A slows intrinsic skin aging and protects from UVB-mediated photodamage: Findings from Randomized Clinical Trials"

Measurements by DermaTOP of skin roughness (wrinkle) - Mean values, standard deviations, treatment comparison on differences to baseline, and time comparison on raw data for serum containing 1% UA

| Parameter | Time | Code | n | Mean Values ± SD | | Treat. Comp. |
|---|---|---|---|---|---|---|
| | | | | Raw Data | Diff. to BL | vs. A |
| Skin Roughness - Ra by DermaTOP [µm] | BL | A | 34 | 22.10 ± 5.80 | -- | -- |
| | | E | 34 | 22.97 ± 5.46 | -- | -- |
| | D15 | A | 32 | 23.84 ± 6.71 | 1.79 ± 4.47 | -- |
| | | E | 32 | 22.14 ± 4.92 | -0.49 ± 3.00 | **0.036** |
| | D57 | A | 34 | 22.62 ± 6.40 | 0.52 ± 3.19 | -- |
| | | E | 33 | 21.64 ± 5.02 | -1.46 ± 3.59 | **0.044** |

**A:** Untreated side of the face**; E:** 1% UA Serum treated side of the face

Ra: average skin roughness;

BL: Baseline; D15: Day 2-weeks of application; D57: 8 weeks of application
