## Supplementary Table 2 for "Topical application of Urolithin A slows intrinsic skin aging and protects from UVB-mediated photodamage: Findings from Randomized Clinical Trials"

Measurements by DermaTOP of skin roughness (wrinkle) - Mean values, standard deviations, treatment comparison on differences to baseline, and time comparison on raw data for day-cream containing 1% UA

| | | | | Mean Values ± SD | | Treat. Comp. |
|---|---|---|---|---|---|---|
| **Parameter** | **Time** | **Code** | **n** | **Raw Data** | **Diff. to BL** | **vs. A** |
| **Skin Roughness - Ra by DermaTOP [µm]** | **BL** | **A** | 33 | 27.41 ± 7.14 | -- | -- |
| | | **B** | 34 | 28.36 ± 8.05 | -- | -- |
| | **D15** | **A** | 34 | 27.22 ± 8.13 | -0.44 ± 4.25 | -- |
| | | **B** | 34 | 24.87 ± 7.33 | -3.33 ± 3.47 | **<0.001** |
| | **D57** | **A** | 35 | 27.41 ± 10.32 | -1.29 ± 5.72 | -- |
| | | **B** | 35 | 24.65 ± 7.61 | -3.81 ± 4.79 | **0.008** |
| **Skin Roughness - Rz by DermaTOP [µm]** | **BL** | **A** | 33 | 111.41 ± 34.99 | -- | -- |
| | | **B** | 34 | 114.43 ± 33.63 | -- | -- |
| | **D15** | **A** | 34 | 111.01 ± 37.95 | -1.49 ± 18.87 | -- |
| | | **B** | 34 | 99.62 ± 29.05 | -13.79 ± 17.49 | **<0.001** |
| | **D57** | **A** | 35 | 111.89 ± 46.78 | -5.85 ± 26.37 | -- |
| | | **B** | 35 | 97.70 ± 31.16 | -17.08 ± 20.99 | **0.009** |

**A:** Untreated side of the face**; B:** 1% UA Treated side of the face

Ra: average skin roughness; Rz: maximum skin roughness

BL: Baseline; D15: Day 2-weeks of application; D57: 8 weeks of application
