## Supplementary Table 1 for "Topical application of Urolithin A slows intrinsic skin aging and protects from UVB-mediated photodamage: Findings from Randomized Clinical Trials"

**Supplementary Table 1: Description of adverse events**

**(*1st Aging Trial*)**

| Event no. | Description of AR | Severity | Relation to product | Onset date | Action taken | Outcome |
|---|---|---|---|---|---|---|
| 1 | Chalazion (right eye) | mild | none | Day 15 | dose not changed | recovered without sequelae |
| 2 | Skin irritation: erythema, papules (right arm) | mild | none (code B) | Day 55 | test product withdrawn | recovered without sequelae |
| 3 | Skin irritation: erythema, papules (left arm) | mild | none (code A) | Day 55 | test product withdrawn | recovered without sequelae |
| 4 | Headache | moderate | none | Day 4 | dose not changed | recovered without sequelae |
| 5 | Covid-19 infection: sore throat, headache, limb pain | mild | none | Day 9 | dose not changed | recovered without sequelae |
| 6 | Skin irritation: erythema, papules, little open wounds, itching (left arm) | mild | none (code B) | Day 51 | dose not changed | recovered without sequelae |
| 7 | Skin irritation: patchy erythema (left arm) | mild | none (code B) | Day 36 | dose not changed | recovered without sequelae |
| 8 | Tissue edema | mild | none (code B) | Day 20 | dose not changed | condition improving |

Of the eight adverse reactions documented in five subjects during the conduct of this study, **none were classified as being serious, and none were deemed related to the test products or placebo.**

(***2**$^{nd}$* ***Aging Trial***)

All adverse reactions that occurred in this study are presented in the following table:

| Event no. | Description of AR | Severity | Relation to product | Onset date | Action taken | Outcome |
|---|---|---|---|---|---|---|
| 1 | Skin irritation: erythema, papules, itching | Mild | None | Day 15 | Dose not changed | Recovered without sequelae |
| 2 | Skin irritation: erythema | Mild | Probable | Day 15 | Dose not changed | Recovered without sequelae |
| 3 | Skin irritation: itching | Mild | Probable | Day 29 | Dose not changed | Recovered without sequelae |
| 4 | Skin irritation: burning | Mild | Probable | Day 1 | Dose not changed | Recovered without sequelae |

Four adverse reactions occurred as varying forms of skin irritation, including erythema, papules, itching and burning. Of the four adverse reactions documented in three subjects during the conduct of this study, all were classified as being of mild severity. In all cases, the dosage of the effected test products remained unchanged and all subjects recovered without sequelae
